# Evaluation of easy-to-implement anti-stress interventions in a series of N-of-1 trials: Study protocol of the Anti-Stress Intervention Among Physicians Study (ASIP)

**DOI:** 10.1101/2024.04.22.24306161

**Authors:** Valentin Max Vetter, Tobias Kurth, Stefan Konigorski

## Abstract

**Background:** Adverse effects of chronically high levels of stress on physical and mental health are well established. In physicians, the effects of elevated stress levels exceed the individual level and include treatment errors and reduced quality of patient-doctor relationships. Breathing and mindfulness-based exercises have been shown to reduce stress and could serve as an immediate and easy-to-implement anti-stress intervention among physicians. Due to the heterogeneity of their effect on stress, we aim to evaluate the intervention effect of performing a short daily breathwork-based or mindfulness-based intervention on the everyday level of perceived stress in physicians in residence in Germany in a series of N-of-1 trials.

**Methods:** Study participants will choose between two short interventions, box breathing, and one guided more complex mindfulness-based breathing exercise. Each participant subsequently will be randomly allocated to a sequence of 1-week intervention (A) and control (B, everyday life) phases. Each N-of-1 trial consists of two two-week cycles (AB or BA), resulting in a total trial duration of 4 weeks (ABAB or BABA). Perceived levels of stress will be assessed daily via the StudyU App on the participant’s smartphone. Additionally, participants will be asked to complete a questionnaire at baseline and three months after completion of the study that contains questions about basic participant characteristics, lifestyle factors, individual living situations, and validated psychological questionnaires. Intervention effects will be estimated by Bayesian multi-level random effects models on the individual and population level.

**Discussion:** This study contributes to the development of short-term solutions to reduce work-related stress for physicians in residence. This is expected to benefit the individual and increase the quality of overall healthcare due to a reduction in treatment errors and an increase in the quality of doctor-patient relationships.

**Trial Registraion:** ClinicalTrials.gov: NCT06368791, first registered April 16, 2024.

## Background

Continuously high levels of psychosocial stress are associated with numerous adverse physical and mental health outcomes [1–3]. Research has consistently demonstrated that physicians represent a demographic notably susceptible to experiencing elevated stress levels over prolonged durations [4]. In a prospective cohort study of 3588 resident physicians in the USA, 45.2% reported symptoms of burnout [5]. Similarly, 48.7% of physicians in surgery in Germany fulfilled the burn-out criteria of the Copenhagen Burnout Inventory (CBI) [6]. In a survey of young German clinicians between their first and sixth year of training, 22% of the participants stated that they had taken medication due to work-related stress [7]. A high prevalence of burnout [8] and a higher stress level compared to the general population were also reported for physicians in general medical practices in Germany [9]. Furthermore, it was repeatedly shown that high levels of stress and burnout in physicians increase the risk for major medical errors [10, 11], and poorer quality of medical care, including poorer doctor-patient relationships and deviation from medical standards [12, 13]. A reduction in stress levels among physicians would, therefore, not only lead to individual benefits with respect to physical and mental health in the medical workforce but also to an improvement in patient care [4] with a potentially positive effect on the health of the general population. Furthermore, reductions in stress and burnout rates have been shown to yield substantial economic advantages [14]. This improvement is partly attributable to lower rates of physician turnover and increased productivity resulting from decreased stress levels [15, 16]. However, sustainable stress reduction is not easy to achieve. For this reason, this study aims to test stress management interventions that are easy to learn and can be performed quickly without the need for special training or additional tools. Special breathing techniques and mindfulness exercises are two approaches that meet these requirements [17, 18]. Box-breathing, also known as “tactical breathing”, is a relaxation technique that decreases heart rate variabililty and, among others, is used by the military and law enforcement to decrease stress levels [18–21]. Breathwork exercises have an impact on a range of symptoms and can lead to a reduction of anxiety, depression, and feelings of anger but also increase the feeling of comfort and relaxation [22, 23]. A recent meta-analysis showed a positive effect of paced breathing on stress perception [24]. Mindfulness interventions vary in type and duration and can positively affect depressive symptoms, anxiety, stress, well-being, and quality of life (meta-analysis in ref. [25]). A meta-analysis of 12 studies reported a positive effect of interventions aimed at reducing stress (including mindfulness-based interventions) [4]. Although the majority of studies in the field focus on the structured 8-week mindfulness-based stress reduction (MBSR) program, there are also studies on shorter intervention periods, including app-based mindfulness interventions [17]. A study that used 10-minute audio recordings of mindfulness exercises over 8 weeks in patients with anxiety disorders showed positive effects compared to a placebo group [26]. A meta-analysis also reported moderate effect sizes of stress-management interventions [27].

Due to interindividual differences and personal preferences, high heterogeneity in intervention effects is expected [28] and has been shown for mindfulness-based interventions [29]. In order to account for this heterogeneity, N-of-1 trials [30] are the gold-standard study design in which every participant applies intervention and non-intervention phases in a cross-over design and continuously assesses the outcome of interest (e.g., daily) over time. After completion of the trial, statistical inference on the intervention effect can be done on the individual level by comparing the outcome in the intervention phases with the outcome in the control phases and on the population level. In the following, we describe a study protocol for a series of N-of-1 trials to evaluate the effect of breathing and mindfulness exercises on stress levels among physicians in residency in Germany.

## Methods/Design

### Study design

In this study, we aim to assess the causal effect of two anti-stress interventions on the daily level of perceived stress. To do so, a series of N-of-1 trials will be conducted. Each trial consists of two phases: (1) eligibility and baseline questionnaire as well as the four-week N-of-1 trial; and (2) a three-months follow-up period after which participants will be asked to fill out a follow-up questionnaire. Since the data is collected anonymously and the participants will be recruited over a period of 2 months, the individual follow-up time between baseline- and follow-up questionnaires can slightly differ between participants. A schematic overview of the study design is shown in Figure 1. Results will be reported following the “CONSORT extension for reporting N-of-1 trials (CENT)” guidelines [31]. The study center is the Institute of Public Health at the Charité – Universitätsmedizin Berlin.

**Figure 1:**
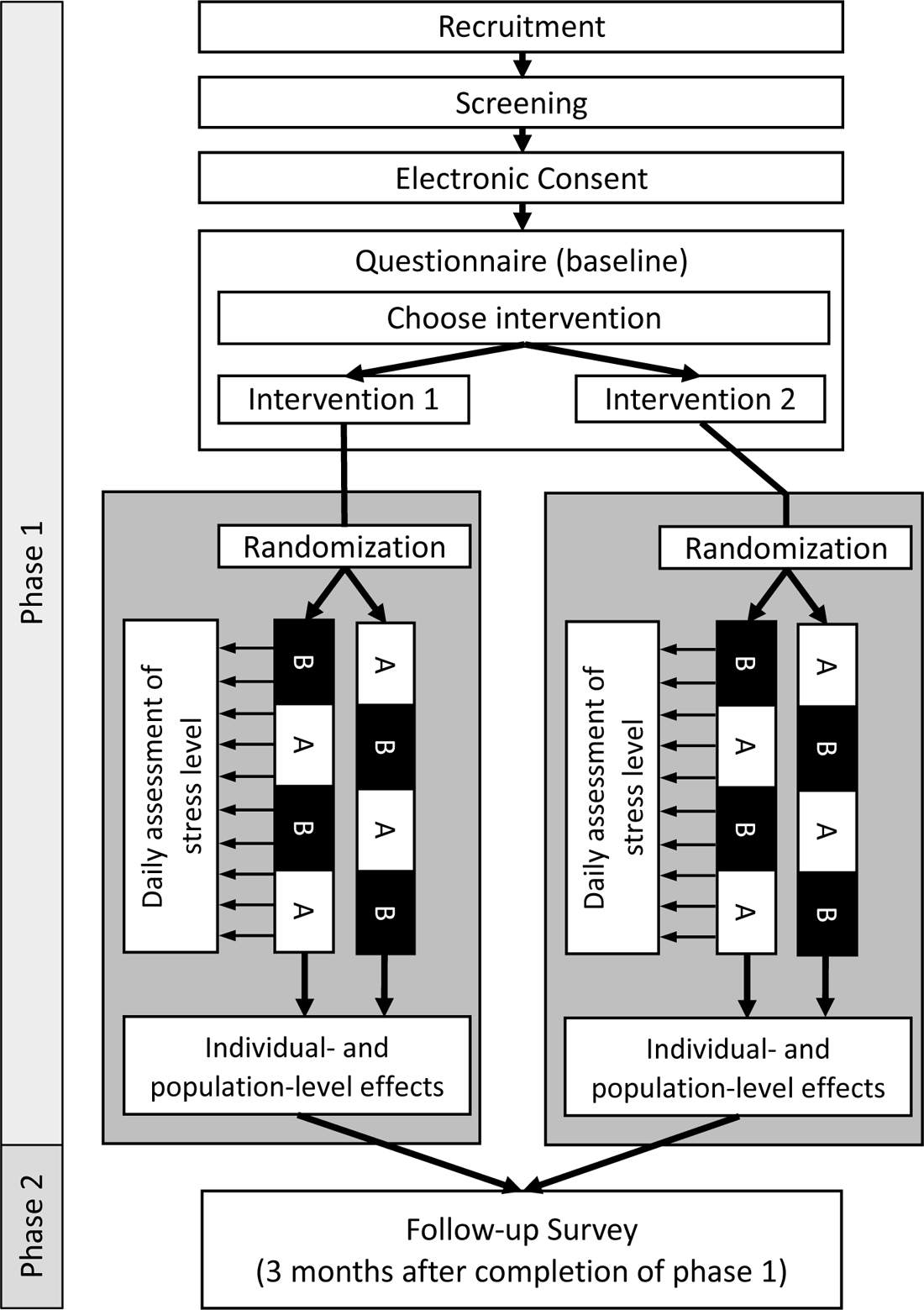
Overview of participant flow and study design. Phase 1: After recruitment and screening for eligibility, participants are asked to give their consent to participate in the study and to fill out the baseline questionnaire. At the end of the questionnaire, participants choose themself which intervention they would like to evaluate. Participants are randomly allocated to one of two possible sequences of intervention (A) and control (B) phases (ABAB or BABA). Phase 2: Three months after completion of Phase 1, participants will be asked to fill out a follow-up questionnaire to assess the sustainability of the intervention.

### Interventions

Participants will be able to choose between two interventions at the beginning of the trial. Both interventions will be provided as video/audio files. The first intervention, a 6-minute box breathing exercise, is used to reduce stress and is employed, for example, by law enforcement and the military to provide effective stress reduction in dangerous situations. Participants are instructed to take some time to relax and sit in a quiet and comforting place. They are then instructed to breathe in for four seconds, hold their breath for four seconds, and breathe out for another four seconds. After holding their breath for four seconds, the next breathing cycle starts. Participants will be provided with a video of a red dot following the outlines of a box at the required speed as a visual aid. The second intervention is a 10-minute guided breathing exercise combining mindfulness meditation with simple body movements and breathwork. Participants are instructed to sit in a quiet room and find an upright position. They are then guided through several short exercises that include light upper body stretching and mindful breathing.

### Participants

The study population is composed of physicians in residence in Germany who work in a hospital or outpatient clinic for at least 9 hours per week. Participants will be recruited via email through mailing lists of professional societies and local training centers of the university hospitals in Germany. Interested participants can sign up for a contact list and will subsequently receive further information as well as a universal access link to the baseline questionnaire per email. Additionally, interested participants can contact the principal investigator via email or phone for further information. Recruitment will be done over the period of two months, and participants are allowed to start with the study as soons as they consent to study participation.

### Inclusion criteria

- Physicians in training in Germany
- Weekly working time in medical activity of at least 9 hours
- Regular access to a smartphone on which the StudyU App can be installed
- Informed consent

### Exclusion criteria

- Age <18 years
- Specialist training already completed
- No clinical activity during the study period (e.g. vacation, research activity, etc.)
- Participation in another intervention study during the study period
- Does not speak German
- Does yoga more than 4 times a month
- Meditates or performs breathing exercises on average more than 4 days per month
- Confirmed or suspected pregnancy
- Presence of a psychiatric disorder
- Presence of cardiovascular disease
- Presence of respiratory or pulmonary disease
- Presence of a neurological disease
- Substance abuse (for example, alcohol, drugs, or other)
- Planned surgery within the next 6 months
- Doctor’s recommendation (or self-assessment) not to perform mindfulness or breathing exercises
- Lack of informed consent
- Employee of the Charité - Universitätsmedizin Berlin (due to data protection reasons, employees of the Charité - Universitätsmedizin Berlin will not be included in this study)

### Procedure

The full study is conducted digitally and only anonymized data will be collected. The anti-stress intervention and guidance throughout the study period, as well as the daily assessment of the participant’s reported outcomes (PROs), will be delivered digitally via the StudyU App [32]. An overview of the study design and participant flow through the trial is shown in Figure 1.

Phase 1: Interested participants will be screened for eligibility and provided with the study information as well as the required consent form via email. If the participants are interested in participating in the study, they can follow a link to the REDCap baseline questionnaire [33] that is provided together with the study documents. After giving consent to participate, the participants are asked to answer the following questions that are used to generate an 8-digit self-generated identification code (SGIC):

- Please enter the first and second letter of your mother’s first name
- Please enter the first and second letter of your place of birth
- Please indicate your sex as stated in your birth certificate.
- Please enter the number of your older (not younger!) siblings.
- Please enter the last digit of your parent’s house number.
- Please enter the last digit of your parent’s postal code.

The SGIC is an adapted version of the approach suggested by Schnell and colleagues [34]. After completion of the baseline questionnaire, participants will be provided with an individual linkage code on the final screen of the questionnaire (which is different from the SGIC). This code can be used (1) by the participants to register for the ASIP study in the StudyU App and (2) by the research team to link the data from the baseline questionnaire with data recorded in the StudyU App. The random allocation of participants to intervention sequence ABAB or BABA (where A is intervention and B is control) is displayed in the app to the participants. Daily push notifications will remind participants to conduct the daily anti-stress intervention during the intervention phase. Additional daily reminders will be sent via push notification to remind reporting the outcome. During control phases, the reminders to document the daily stress level will be displayed on the participant’s phone via push notification.

If the minimally required number of PROs was documented, participants will be given access to an automatically generated report with the results of their collected data within the StudyU App after the trial is successfully completed.

Phase 2: Three months after completion of phase 1, participants will be asked to fill out a follow-up questionnaire. At the beginning of this questionnaire, they will be asked to provide their SGIC, which will be used to link it to the baseline questionnaire. The follow-up questionnaire includes the same items as the baseline questionnaire but additionally asks the participants to report on adverse events and if and how the intervention was implemented in everyday life after study completion.

### Outcomes

Basic characteristics and additional information about participants, will be assessed through digital surveys using the REDCap [33] at baseline and three months after completion of phase 1. Additionally, as part of the N-of-1 trial (Phase 1, Figure 1), daily PROs will be collected with the StudyU App [32].

Primary Outcomes: Participants will be asked to answer the following two questions daily in the StudyU App on a visual analog scale from 1 (“not stressed at all”) to 10 (“extremely stressed out”):

– “Overall, how stressful was your day?”
– “Which level of stress do you expect for the following day?”

Participants will be able to answer these questions after 4 p.m. each day and will be reminded with push notifications if the PRO is not documented. It should be noted that the second question targets the anticipated stress on the following day independently of the intervention effect. This is naturally the case in the control phase, and we assume that participants will also not include the anticipated intervention effect in their anticipated rating of the stress on the following day so that the answers in both phases can be compared.

Additional secondary outcomes are the level of agreement between expected and actually perceived levels of stress, adherence to study protocol, and successful completion of the study (defined as reporting a minimally required number of PROs during phase 1 of the study).

### Effect Modifiers and Additional Variables

To characterize the study participants and also exploratively investigate potential effect measure modifiers, additional variables are assessed as part of the self-administered surveys at baseline and follow-up. Participants are asked to provide information about age, sex, place of residence, place of work (hospital vs. outpatient/ambulatory setting),, regular work hours per week, lifestyle variables (sport, alcohol, smoking), as well as the individual living situation, relationship status and number of children in the household. Additionally, selected questions from the validated German versions of the following standardized and free-to-use questionnaires are used to assess additional psychological variables:

– Copenhagen Psychosocial Questionnaire (COPSOQ, work-life conflicts) [35, 36]
– Copenhagen Burnout Inventory, Personal Burnout [35]
– Effort-reward imbalance questionnaire (ERI) [37, 38]
– ERI Overcommitment [37, 38]
– Cohen’s Perceived Stress Scale [39, 40]
– Satisfaction with life scale (SWLS) [41]
– System Usability Scale (SUS)

### Data management

Data will be collected anonymously, i.e., no data is collected that allows to connect the collected data with identifying information at any point in time. Data collected through the REDCap survey at baseline and the data collected in the StudyU App will be linked using a randomly allocated linkage code automatically and digitally provided at the end of the baseline survey. This code is needed to get access to the ASIP study in the StudyU App and will be used to link data from both databases (REDCap and StudyU) after study completion. Data collected with the REDCap survey at baseline is linked with data collected in the follow-up REDCap survey three months after completion of phase 1 by the SGICs that are only known to the participants. Two months after the linkage of the baseline and follow-up questionnaire at the end of the study, the SGICs will be deleted from the data.

Anonymous data from the REDCap surveys will be stored on protected servers of the Charité – Universitätsmedizin Berlin. Anonymous data collected through the StudyU App is saved securely on a backend hosted locally by the Hasso-Plattner-Institute in Potsdam, Germany. Until the SGICs are deleted, all data will be stored password-protected, and only the principal investigator and nominated research staff will have access to the data. After completion of the study and deletion of the SGIC, parts of the data will be published in a public research data repository. Data collected in the StudyU App will be published in the StudyU repository and made openly available.

### Estimand

In this study, the level of stress is assessed daily between 4 p.m. and the end of the day and should only be answered after the intervention is completed. Participants are instructed to perform the anti-stress intervention via push notification on their mobile phone at 10 a.m. on each day of the intervention phase but are free to perform the intervention whenever they choose. There might not be full adherence to the intervention suggestions and the adherence is not assessed. Hence, only information about the availability of the intervention through the pre-specified intervention sequence is available.

Consequently, in this study, we are targeting the causal effect of the availability of each of the two anti-stress interventions among participants who chose the respective intervention prior to the study. In particular, we are interested in the effect of “always performing the intervention” compared to “never performing the intervention..” Under the assumption of full adherence, this estimated causal effect of intervention availability is equal to the effect of the intervention itself. We assume no carryover effects, that the treatment effect is constant over time and that there is no effect of time-varying variables throughout the study period. We will calculate individual effects as well as the average effect across participants.

### Sample size

Sample size was calculated for the population effect of the two investigated anti-stress interventions on the daily level of perceived stress assessed on an analog scale between 1 and 10. To calculate the sample size, the approach described by Yang and colleagues [42] was employed, which uses a linear mixed model to estimate sample size in a series of N-of-1 trials.

The ShinyApp web application (https://jiabeiyang.shinyapps.io/SampleSizeNof1/) provided by the authors was used, and a fixed intercept and common slope model were assumed. Calculations were done based on a homogeneous residual standard error of 2.41, which is in line with results published by Lesage and colleagues [43]. Based on results published by Naughton and Johnston [44], an autocorrelation between PROs at two adjacent time points (i.e., days) of 0.8 was assumed. For this sample size calculation, an alternating sequence was assumed (ABAB or BABA). Due to the novelty of this study, an estimation of the expected effect size is challenging. Therefore, we conservatively assumed an intervention effect of a standardized mean difference of at least 0.3 points between intervention and control phases, which is 50% of the effect size reported in a meta-analysis of ten studies investigating the effect of mindfulness-based intervention on stress over a longer time period in future health professionals [45]. Setting the level of statistical significance as α = 5% and assuming documentation of PROs on at least 4 days per week, a power of 80% will be reached with six participants per sequence(ABAB or BABA), yielding n=12 participants in total for each of the two investigated interventions. Under an assumption of a dropout rate of 30% (so that the full trial data cannot be used), n=17 participants need to be recruited per intervention (n=34 participants in total).

### Statistical analysis

To estimate the estimand of interest described above for each intervention individually, both interventions will be seperately analyzed in comparison to the everyday-life control. On the individual level, intervention effects will be estimated using Bayesian repeated-measure models which will include a first-order autoregressive error structure. Autocorrelation of daily stress was observed before [44] and will be considered during the data analysis. For population-level effect estimation, the data from all individual N-of-1 trials will be analyzed together. A Bayesian multilevel repeated-measures model will be used to estimate the posterior distribution of the population-level average intervention effect as well as the within- and between-participant variance. Similarly to the individual-level estimates, the dependency of responses over time will be modeled by a first-order autoregressive error structure. Statistical analyses will be conducted with JAGS [46] and the statistical software package R [47] employing the Markov Chain Monte Carlo (MCMC) method. We will perform an intention-to-treat analysis.

As a secondary analysis, we will perform a joint analysis of both interventions to estimate the effect of the availability of an anti-stress intervention among the participants that chose the respective intervention. Additionally, we will perform an exploratory analysis of effect measure modifiers using Bayesian models.

### Monitoring and safety

Since the intensity of the breathwork and mindfulness exercises is rather low, we estimate the risk for adverse outcomes to be very low. Nevertheless, in accordance with other studies in the field, a number of exclusion criteria were defined to further minimize the risk of unwanted negative outcomes. As part of the follow-up questionnaire three months after study completion, participants will be asked if and which adverse events were experienced due to the intervention.

## Discussion

The Anti-Stress Intervention Among Physicians (ASIP) study aims to evaluate the effect of two anti-stress interventions on the daily level of perceived stress in physicians in residence in Germany. To assess the efficacy of these interventions, a series of N-of-1 trials will be conducted. Participants can choose between box-breathing and a mindfulness-based breathing exercise, thereby studying the effect of their preferred intervention, and will randomly be allocated to a sequence of intervention (A) and control (B) phases (resulting in sequence ABAB or BABA). The study participants will report their perceived stress levels daily through a mobile application. Bayesian analyses will be conducted to estimate the intervention effect and effect of potential effect modifiers such as age, sex, work experience, working hours per week, and others will be investigated.

Strengths of this study include its design (series of N-of-1 trials), which enables the examination of intervention effects on the individual and the population level. The first is of special interest in the case of stress-reducing interventions, as the individual effect is the one that we ultimately want to achieve. Also, for the estimation of effects on the group level, due to repeated assessment of the PROs, a smaller sample size compared to traditional RCTs is sufficient to reach the same statistical power. Both interventions are short and easy to conduct (6 and 10 minutes) and, therefore, easy to implement into everyday life. This potentially increases adherence to the study protocol and long-term use (if proven successful). Additionally, each participant will be given access to their individual N-of-1 trial results. This could potentially motivate participants to use the intervention in their everyday lives even after completion of the study.

We want to point out several limitations to this study: First, the control phases are generally routine in everyday life. Therefore, no placebo control is available, which is a frequent problem in studies evaluating anti-stress interventions [17, 26]. However, since we are ultimately interested in the evaluation of the total intervention effect, it could be argued that, although some part of the observed effects might be attributable to the placebo effect, this is acceptable as the intervention itself only introduces a limited burden to the conducting person and adverse outcomes are unlikely. Second, protocol adherence and dropout are expected to be a challenge in this study. Therefore, we aim to limit deviation from the study protocol due to individual guidance through the study procedure as well as the delivery of the intervention and collection of the daily PRO through a mobile application. Furthermore, conservative sample size calculations included a dropout rate of 30%, and the pre-specified power of 80% for the estimation of group effects is reached if PRO are reported on 4 of the 7 days per intervention or control phase. Therefore, a robust study design is in place to deal with incomplete data reporting and high drop-out rates.

## Conclusion

High levels of stress are known to be responsible for numerous adverse health outcomes. In the case of physicians, it results in negative outcomes exceeding the individual negative health effects due to increased treatment errors and decreased quality of the patient-doctor relationship. Therefore, if proven effective, the anti-stress interventions investigated in this study could potentially be useful in the sustainable decrease of stress in physicians and thereby improve individual and population-level health.

## Data Availability

Anonymously collected data will be made in part publicly available via research data repositories after study completion.

## Acknowledgments

The authors would like to thank Ellen Zitzmann for her valuable input and help in developing the mindfulness-based breathing exercise and recording the intervention audio files. In addition, the authors thank Marco Piccininni for his notes on the design and the statistical analysis plan of this study.

Ethics approval and consent to participate

All participants provided informed consent, and the study was conducted in accordance with the Declaration of Helsinki and approved by the Ethics Committee of the Charité-Universitätsmedizin Berlin— approval number EA4/260/23.

## Authors contributions

VMV: initial idea; precise planning and conception of the study; implementation of questionnaires; statistical analysis; drafting the manuscript; has given final approval of the version to be published. TK: conception and design of the study; supervision; revising the manuscript critically for important intellectual content; has given final approval of the version to be published. SK: conception and design of the study; supervision; statistical analysis; revising the manuscript critically for important intellectual content; has given final approval of the version to be published.

